# Overdoses with Xylazine and Fentanyl Recorded in Pennsylvania’s Overdose Information Network: An Analysis of Law Enforcement/First Responder-reported Overdose Response

**DOI:** 10.1101/2024.08.29.24312792

**Authors:** Manuel Cano, David T. Zhu, Yesenia Aponte-Meléndez, Pedro Mateu-Gelabert, Alex S. Bennett

## Abstract

This study explored whether law enforcement/first responder-reported fentanyl overdose response actions (such as administration of the opioid overdose reversal agent naloxone) differed between overdoses in which xylazine was, versus was not, suspected to be co-involved. Data were drawn from the Pennsylvania State Police’s Overdose Information Network (ODIN) for 11,478 suspected fentanyl-involved overdoses, 137 reportedly co-involving xylazine, recorded across Pennsylvania, excluding Philadelphia, January 2018-January 16, 2025. We used relative frequencies, Fisher’s exact tests, and binomial logistic regression to compare first responders’ overdose response actions in suspected fentanyl overdoses cases in which xylazine was, versus was not, reportedly co-involved. Naloxone was administered at the scene of 46.0% of the overdoses reportedly involving fentanyl and xylazine, vs. 67.3% of the reported fentanyl-no-xylazine overdoses. Multivariable regression results (among the suspected fentanyl overdoses in ODIN, adjusting for age, sex, race/ethnicity, year, county rurality, and other drugs suspected to be involved) indicated that suspected xylazine co-involvement was associated with 60% lower odds of naloxone administration (Adjusted Odds Ratio, 0.40; 95% Confidence Interval, 0.28-0.57). Observed differences in overdose response based on suspected xylazine co-involvement support the importance of equipping first responders with the tools and training to recognize/manage the distinct challenges of xylazine-fentanyl-involved overdose.

## Introduction

Xylazine, a veterinary non-opiate sedative and analgesic, is increasingly detected in overdose deaths in the United States (US), usually accompanied by fentanyl (Spencer et al., 2023). Philadelphia, Pennsylvania is considered the earliest epicenter of xylazine-adulterated drugs in the mainland US (Friedman et al., 2022; Zhu, 2023), and xylazine-related overdose deaths have primarily been concentrated in the northeastern US (Spencer et al., 2023). Nonetheless, recent studies suggest that xylazine’s presence in illicit drug supplies may be spreading throughout the US as well as several other nations (Friedman et al., 2022; Bradford et al., 2024; Bufanda et al., 2025; Cano et al., 2024; Copeland et al., 2024; Delcher et al., 2024; Friedman et al., 2024; Thomas et al., 2024; Wu et al., 2024).

The potential impacts of xylazine on overdose outcomes are unclear (D’Orazio et al., 2023; Hoffman, 2023). Research with rodents suggests xylazine may increase risk of opioid overdose fatality (Choi et al., 2023). In contrast, an analysis of opioid overdose patients in nine US emergency departments documented less cardiac arrest and coma in xylazine-positive cases compared to non-xylazine cases (Love et al., 2023). Regardless of xylazine’s specific role in overdose risk, polysubstance overdoses co-involving xylazine and fentanyl present additional challenges for clinical management and public health strategies (Gupta et al., 2022).

One of these challenges is related to what steps bystanders and first-responders can take to help reverse potential xylazine-fentanyl-related overdoses; bystander and first-responder actions are crucial because fentanyl’s rapid and potent effects narrow the window of opportunity to prevent death or long-term damage (del Pozo, 2022; Fairbairn et al., 2017). For overdoses involving opioids, one of the primary recommendations is for bystanders and first responders to administer the opioid overdose reversal medication naloxone. Since xylazine is not an opioid, naloxone does not directly address xylazine’s effects (Gupta et al., 2022), but naloxone is still recommended as a critical tool to save lives in cases of xylazine-involved overdose (Zhu, 2023; Gupta et al., 2023; Zagorski et al., 2023) because opioids like fentanyl are nearly always co-involved (reported in 99% of xylazine-related overdose fatalities nationwide; Spencer et al., 2023).

Some bystanders and non-medical first responders may be deterred from administering naloxone if xylazine is believed to be involved in the overdose, based on media headlines referring to xylazine as “resistant” to naloxone (e.g., Talbert, 2024). At the same time, other concerns have been documented regarding bystanders or first responders potentially administering more doses of naloxone than needed in cases involving xylazine, which may worsen withdrawal symptoms (Reed et al., 2025; Quijano et al., 2023). Even after a naloxone dose may have restored a person’s breathing (reversing effects of the fentanyl involved in the overdose), the person may still appear unresponsive due to xylazine’s sedative properties (Bufanda et al., 2025), sometimes leading responders to continue rapidly administering more naloxone (Quijano et al., 2023).

A better understanding of first responders’ rates of naloxone administration in overdoses potentially involving xylazine and fentanyl can help identify opportunities to strengthen overdose prevention and response efforts. Prior studies have examined rates of naloxone receipt in *fatal* xylazine-involved overdoses; an analysis of data from Tennessee indicated that naloxone had been administered in 34% of xylazine-involved overdose deaths from 2019-2022 (Korona-Bailey et al., 2023), and an analysis of deaths in 21 US jurisdictions found that naloxone had been administered in 24% of the fentanyl overdoses with or without xylazine detected (Kariisa et al., 2023). These studies focused on fatal overdoses only, and as such, naloxone administration rates reflected in these studies may not likely generalize to overdoses overall, including nonfatal events.

Therefore, the present study examines naloxone administration at the scene of both fatal and non-fatal fentanyl overdoses with and without xylazine reportedly involved, leveraging data from a centralized repository of overdose events recorded by law-enforcement and other participating first-responders across Pennsylvania, excluding Philadelphia. Whereas studies of naloxone administration in xylazine-involved deaths (Kariisa et al., 2023; Korona-Bailey et al., 2023) offer data on drug positivity obtained from postmortem toxicology testing, the present study focuses on *first responders’ assessment/report* of xylazine involvement in overdose, based on information available to them at the scene and time of overdose (from sources such as drug field testing, investigating the scene, and interviewing those present). First-responders’ report of suspected xylazine involvement, whether over- or under-estimating true xylazine positivity, is of interest in this study because first responders’ actions (e.g., whether to administer naloxone, if available) are based on their information/perceptions *at the time of* overdose response, rather than what full toxicology testing would have determined.

## Methods

### Study Design and Sample

We analyzed data from the publicly available version of the Pennsylvania State Police’s Overdose Information Network (ODIN; Pennsylvania State Police, 2024), a repository recording overdoses attended by participating law enforcement agencies (or voluntarily reported by other first responders such as fire or emergency medical services). ODIN was formed by the Pennsylvania State Police and the Liberty Mid-Atlantic High Intensity Drug Trafficking Area to provide “real-time information to aid in drug and overdose investigations,” including data on “fatal and non-fatal drug overdoses, naloxone administrations and identifying markings found on drug packaging” (Rhoads, 2019, p. 2). ODIN is used to provide “overdose spike notifications” to local and state leaders in consultation with the Pennsylvania Department of Health (SB 2158, 2022). ODIN does not represent all Pennsylvania overdoses, nor all law enforcement-attended overdoses (not including, for example, overdoses in which 911 was never contacted or in which the responding personnel were not ODIN reporters). Nonetheless, ODIN has been used in prior analyses of Pennsylvania overdoses (Barboza & Angulski, 2020; Cano et al., 2025; Homes & King, 2023; Holmes et al., 2022; King et al., 2021) due to the critical information offered regarding drugs suspected in overdoses, naloxone administration, and overdose survival or fatality.

The study’s primary analytic sample comprised 11,478 suspected fentanyl-involved overdoses, each with a unique victim ID/incident ID combination, and 137 of these overdoses (1.2%) also included xylazine as a suspected drug. Additional analytic subsamples used in the study included: a) the subsample of 7,550 suspected fentanyl-involved overdoses (62 reportedly co-involving xylazine) in which naloxone was reportedly administered; and b) the subsample of 6,579 suspected fentanyl-involved overdoses (42 reportedly co-involving xylazine) in which naloxone was reportedly administered and the person survived. In Supplemental Table S1, we summarize the process followed to identify unique records and merge data regarding multiple drugs suspected in each overdose. Since the Philadelphia Police Department was not included as one of the 752 ODIN-reporting agencies (as of the study date), we limited our study to overdoses outside of Philadelphia County (thereby excluding the 80 [less than 1%] fentanyl-involved overdoses from Philadelphia County). The study data included overdoses recorded between January 1, 2018-January 16, 2025 (from the ODIN dataset updated on January 16, 2025). The study was classified as “exempt” by the Arizona State University Institutional Review Board based on Federal Regulation 45CFR46 (4).

### Measures

All measures in ODIN are based on the report of first responders such as law-enforcement; for example, “suspected drugs” are determined by investigation of evidence at the scene, information gleaned from interviews at the scene, and/or field testing (Barboza & Angulski, 2020). In the present study, our exposure of interest was a binary indicator (yes/no) for whether xylazine was recorded as a “suspected” drug. We examined demographics (sex, age category, and race/ethnicity), other drugs recorded as “suspected drugs” involved in the overdose (binary indicators for heroin, carfentanil, other synthetic opioids or fentanyl analogs, pharmaceutical opioids, methamphetamine, cocaine, benzodiazepines, and alcohol), overdose survival (yes/no/unknown, as reported in ODIN by the responding agency), rurality of the county of occurrence (metro, or non-metro, based on the 2013 National Center for Health Statistics Urban-Rural Classification Scheme for Counties), and year (with the years 2024 and 2025 combined because only two weeks of data from 2025 were available at the time of data analysis). We focused on three outcome measures: 1) whether naloxone was administered at the scene (yes/no); 2) the quantity of naloxone doses reportedly administered at the scene (in categories of one, two, or three or more, applicable only for cases in which naloxone was administered); and 3) responders’/patients’ actions following naloxone administration, in cases in which naloxone was reportedly administered and the patient survived, with the following mutually-exclusive and exhaustive options available: arrest, release/verbally refer to treatment, transport to treatment, refuse transport, transport to hospital conscious, transport to hospital unconscious, and other/unknown.

### Analyses

First, we summarized characteristics of suspected fentanyl overdoses with vs. without xylazine reportedly involved, calculating descriptive statistics for demographics, drug involvement, survival, and naloxone administration, using Fisher’s exact tests for comparisons (due to small numbers in some categories). Next, we used binomial logistic regression to test the association between first responders’ report of *suspected xylazine involvement* (in fentanyl overdoses recorded in ODIN) and *administration of naloxone* at the scene (yes/no), adjusting for sex, age, race/ethnicity, county rurality, year, and other “suspected” drugs involved, with results expressed using adjusted odds ratios (AORs) and 95% Confidence Intervals (CIs) as well as average marginal effects (AMEs; i.e., the expected change in naloxone administration if xylazine is suspected to be involved in the fentanyl-related overdose). Supplemental analyses repeated the regression model with the following changes: a) changed the analytic sample from suspected fentanyl overdoses only to overdoses involving any type of opioid; and b) limited the timeframe to January 2022-January 16, 2025, since xylazine was rarely reported in ODIN prior to 2022.

Next, we focused on the subset of suspected fentanyl overdoses recorded in ODIN in which naloxone was reportedly administered at the scene. In this subset of overdoses, we used Fisher’s exact tests to compare the number of naloxone doses administered in suspected fentanyl overdoses with vs. without xylazine reportedly involved. We added a supplemental analysis to compare the proportions of fatal and nonfatal suspected fentanyl overdoses in each naloxone dose category to determine whether fatality might have been more common in any dose category. Finally, we focused on the subset of suspected fentanyl overdoses recorded in ODIN in which naloxone was reportedly administered at the scene and the patient survived, using Fisher’s exact tests to compare first responders’/patients’ reported action after naloxone was administered. All analyses were completed in Stata/MP 18.0. The regression analysis used listwise deletion for missing data (with a total of 4.2% missing/unknown on all measures combined), and in descriptive analyses of quantity of naloxone doses, we eliminated the 147 (1.3%) overdoses in which the number of naloxone doses was not consistent between different administering agencies’ report of the same incident/victim ID.

## Results

ODIN-recorded suspected fentanyl overdoses, either with (n=137) or without (n=11,341) xylazine suspected to be involved, were concentrated among men, in ages 30-39, among Non-Hispanic (NH) White individuals, and in Allegheny and Montgomery counties (Supplemental Tables S2-3). As depicted in Figure 1, several drugs were reported in significantly larger proportions of overdoses in the suspected fentanyl-xylazine group than the fentanyl-no-xylazine group: carfentanil (10.2% vs. 2.2%; *p*<0.001); other synthetic opioids/fentanyl analogs (20.4% vs. 7.6%; *p*<0.001); methamphetamine (20.4% vs. 7.0%; *p*<0.001); and benzodiazepines (5.1% vs. 2.4%; *p*=0.046). Conversely, heroin was reported in less than half (46.0%) of the overdoses in the xylazine group versus two thirds (66.3%) in the no-xylazine group (*p*<0.001). Naloxone was administered in 46.0% of the cases reportedly involving fentanyl-with-xylazine, versus 67.3% of the fentanyl-no-xylazine cases (*p*<0.001), and death was the outcome of 54.7% of the cases reportedly involving fentanyl-with-xylazine, compared to 27.0% of the fentanyl-no-xylazine cases (*p*<0.001).

**Figure 1.**
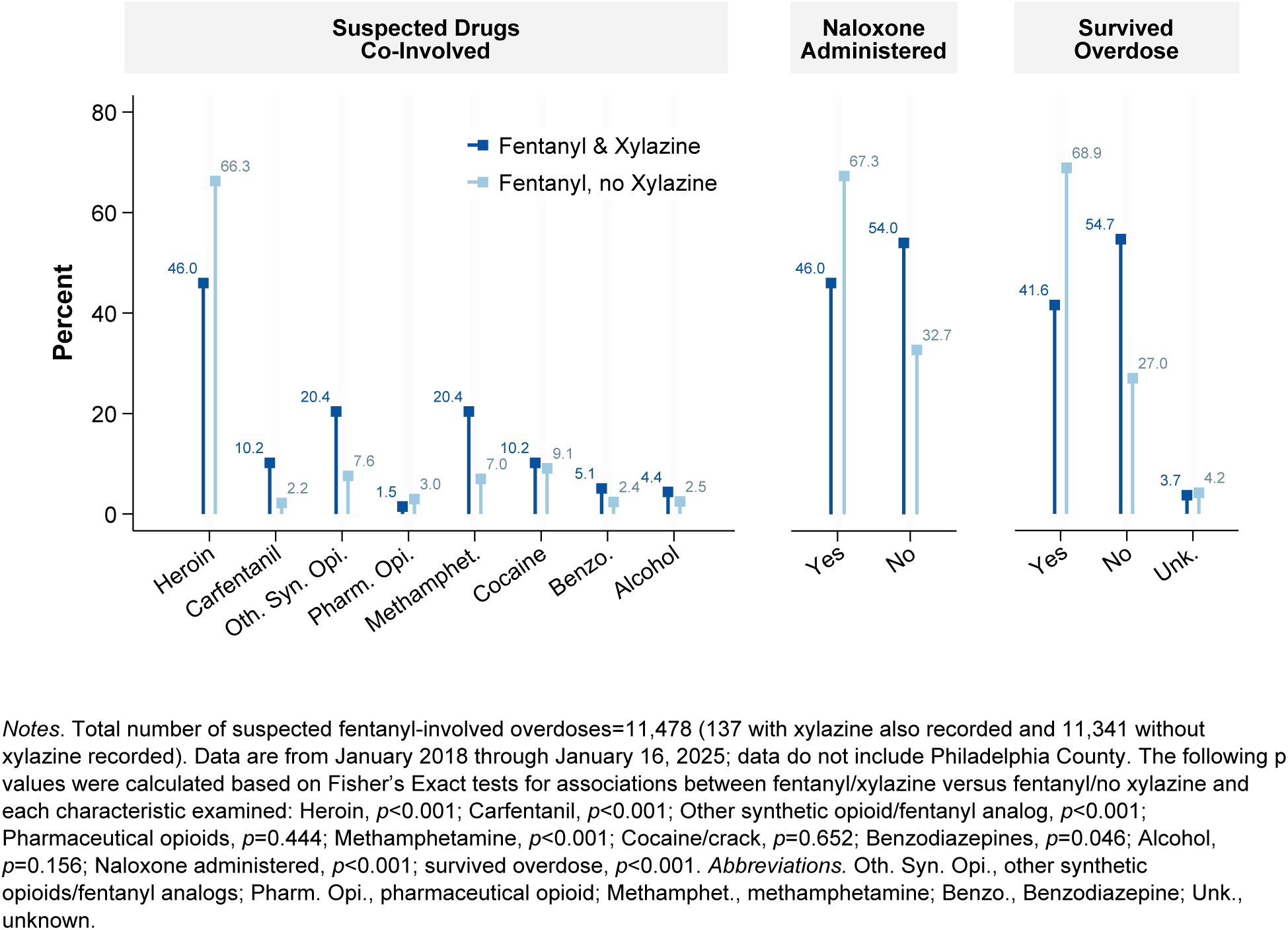
Characteristics of overdose events reportedly involving fentanyl and xylazine versus fentanyl and no xylazine, as recorded in the Pennsylvania Overdose Information Network, January 2018-January 16, 2025.

In the multivariable regression model (Table 1), even after adjusting for age, sex, race/ethnicity, year, county rurality, and suspected involvement of other drugs, suspected xylazine involvement was associated with 60% lower odds of naloxone administration (AOR 0.40; 95% CI, 0.28-0.57; or 19 percent lower probability [AME −0.19; 95% CI −0.27,−0.12]) among the suspected fentanyl overdoses recorded in ODIN. Results from supplemental analyses were similar (Supplemental Table S4) when broadening the analytic sample to overdoses involving any type of opioid (instead of fentanyl specifically) and when restricting the analysis time frame to the years xylazine was most frequently reported in ODIN (2022-2025).

**Table 1.** Results of binomial logistic regression predicting naloxone administration, based on suspected xylazine involvement and selected covariates, among suspected fentanyl-involved overdoses reported in the Pennsylvania Overdose Information Network, January 2018-January 2025.

| Outcome: Naloxone Administration<br>n=10,991 |  |
| --- | --- |
|  | AOR [95% CI] |
| Xylazine co-involved | **0.40\*\*\*** [0.28, 0.57] |
| Age, years |  |
| 0-9 | 0.96 [0.43, 2.15] |
| 10-19 | **1.51\*** [1.02, 2.24] |
| 20-29 | 1.10 [0.99, 1.23] |
| 30-39 | 1.00 [reference] |
| 40-49 | **0.85\*\*** [0.76, 0.95] |
| 50-59 | **0.76\*\*\*** [0.66, 0.87] |
| 60-69 | **0.69\*\*\*** [0.57, 0.85] |
| 70+ | 0.90 [0.48, 1.70] |
| Sex |  |
| Male | 1.00 [reference] |
| Female | 1.00 [0.91, 1.09] |
| Race/ethnicity |  |
| NH White | 1.00 [reference] |
| NH Black | **1.57\*\*\*** [1.35, 1.83] |
| Hispanic | **1.76\*\*\*** [1.46, 2.13] |
| NH AI/AN | 0.20 [0.04, 1.01] |
| NH Asian | 1.13 [0.47, 2.71] |
| Other drugs co-involved |  |
| Heroin | **1.32\*\*\*** [1.20, 1.45] |
| Carfentanil | 1.03 [0.78, 1.38] |
| Other synthetic opioid/fentanyl analog | **1.20\*** [1.02, 1.41] |
| Pharmaceutical opioids | 0.93 [0.72, 1.18] |
| Methamphetamine | **0.50\*\*\*** [0.43, 0.58] |
| Cocaine/crack | **0.48\*\*\*** [0.42, 0.55] |
| Benzodiazepines | **0.61\*\*\*** [0.47, 0.79] |
| Alcohol | 0.79 [0.61, 1.01] |
| Metropolitan Status |  |
| Non-Metro | 1.00 [reference] |
| Metro | **1.25\*\*** [1.09, 1.43] |
| Year |  |
| 2018 | 1.00 [reference] |
| 2019 | 1.12 [0.91, 1.37] |
| 2020 | 1.19 [0.99, 1.44] |
| 2021 | 1.11 [0.92, 1.34] |
| 2022 | **1.33\*\*** [1.10, 1.61] |
| 2023 | **1.60\*\*\*** [1.34, 1.92] |
| 2024/2025 | **1.48\*\*\*** [1.23, 1.79] |
| AME [95% CI] |  |
| Average Marginal Effect (AME), xylazine co-involvement, [95% CI] | **-0.19\*\*\*** [-0.27, -0.12] |

*Note.* Data are from January 2018 through January 16, 2025; data do not include Philadelphia County. Naloxone administration denotes if naloxone was administered to the individual who overdosed. Average Marginal Effects (AME) denote the proportional change in naloxone administration when xylazine changes from "not suspected to be involved" to "suspected to be involved" in the overdose. Estimates in bold highlight statistical significance (two-tailed): \* $p < .05$ , \*\* $p < .01$ , \*\*\* $p < .001$ . *Abbreviations:* NH, Non-Hispanic; AI/AN, American Indian/Alaska Native; AOR, adjusted odds ratio; CI, confidence interval.

As depicted in Figure 2, within the subset of suspected fentanyl overdose events in which naloxone was administered, the quantity of naloxone doses differed by suspected xylazine co-involvement (*p*=0.012). A single dose of naloxone was administered in 56.3% of the suspected fentanyl-no-xylazine overdoses with naloxone administered, compared to 41.9% of the suspected fentanyl-xylazine overdoses. Three or more doses were recorded in one in ten (10.1%) of the suspected fentanyl-no-xylazine overdoses with naloxone administered but two in ten (21.0%) of the suspected fentanyl-xylazine overdoses. In supplemental analyses (Supplemental Table S5), we compared the proportion of deaths in ODIN-recorded suspected fentanyl overdose events based on naloxone dose count (one, two, or three or more) and did not observe any statistically significant difference in death proportions between these three dose groups (*p*=0.235).

**Figure 2.**
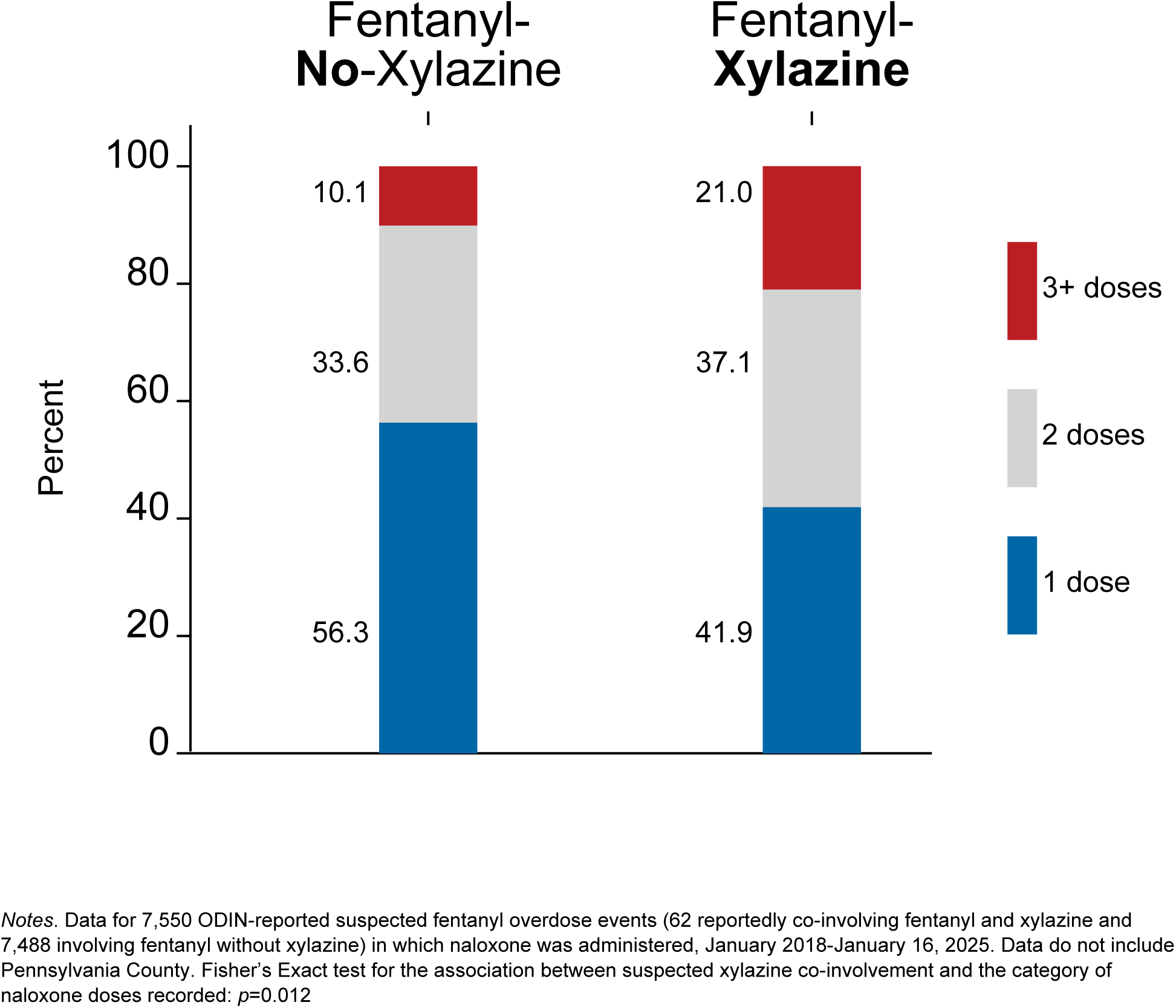
Number of naloxone doses administered in fentanyl-related overdose events in which naloxone was administered, by suspected xylazine co-involvement, as recorded in the Pennsylvania Overdose Information Network, January 2018-January 16, 2025.

Figure 3 depicts post-naloxone first responder/patient actions in the subset of suspected fentanyl overdoses in which naloxone was administered and the patient survived. In this subsample (42 suspected fentanyl-xylazine overdose events and 6,537 suspected fentanyl-no-xylazine events), the share of patients arrested was higher in the suspected fentanyl-xylazine group (4.8%) than the fentanyl-no-xylazine group (1.8%). The proportion of patients transported to the hospital unconscious was also higher in the suspected fentanyl-xylazine group (14.3%) than the fentanyl-no-xylazine group (5.6%). Conversely, the proportion of patients who reportedly refused transport was lower in the suspected fentanyl-xylazine group (4.8%) than the fentanyl-no-xylazine group (23.2%).

**Figure 3.**
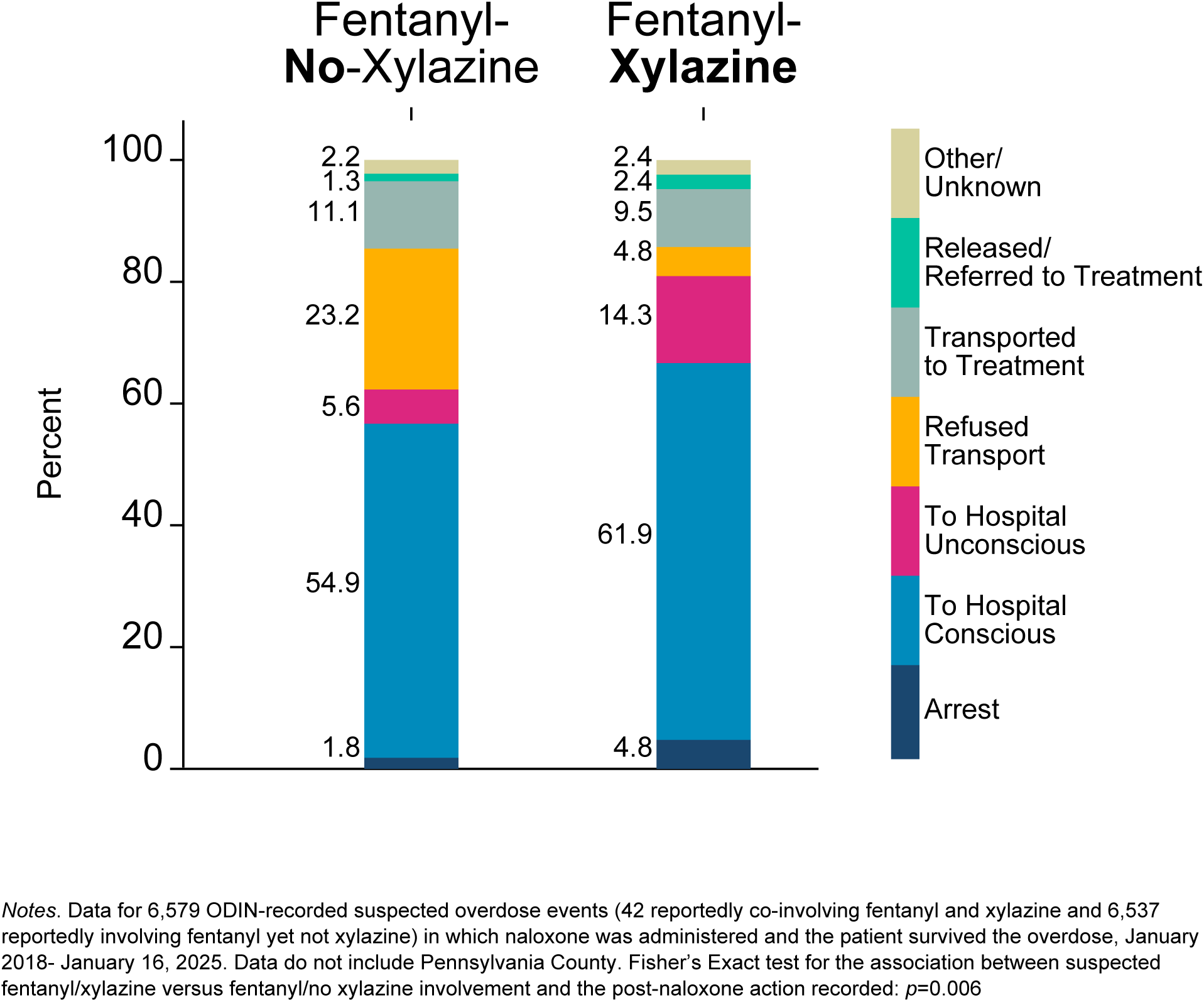
Post naloxone-administration actions in fentanyl-related overdose events in which naloxone was administered and the patient survived, by suspected xylazine co-involvement, as recorded in the Pennsylvania Overdose Information Network, January 2018-January 16, 2025.

## Discussion

This study examined Pennsylvania law enforcement/first responders’ reported overdose response actions in fentanyl-involved overdose events with (vs. without) xylazine reported as a suspected co-involved drug. Study findings highlighted several differences in overdose response actions (as well as overdose characteristics) based on suspected xylazine co-involvement. These differences are detailed in the paragraphs that follow. The role of law enforcement officers in overdose response (Del Pozzo, 2022; Doe Simpkins et al., 2022) is complex for various reasons; for instance, previous evidence indicates that the risk of arrest may deter some individuals from calling 911 when witnessing an overdose (Koester et al., 2017; Wagner et al., 2019).

Nonetheless, Pennsylvania police have been permitted to administer naloxone (and required to complete online training on the topic) since 2014, and a 2018 study found that 91% of surveyed Pennsylvania law enforcement officers reported they had access to naloxone, and 73% indicated they were usually the first on the scene of an overdose (Jacoby et al., 2020).

### Naloxone administration rate differences based on suspected xylazine co-involvement

The study’s first finding was that naloxone was administered at the scene of overdose in a lower share of the overdoses suspected to involve fentanyl with xylazine (46%), compared to overdoses suspected to involve fentanyl but no xylazine (67%). Even after adjusting for other drugs suspected to be co-involved in the overdose, as well as patient demographic characteristics, county rurality, and year, suspected xylazine co-involvement was significantly associated with lower probability of naloxone administration at the scene of the overdose. All the cases included in the present study reportedly involved fentanyl, for which timely naloxone administration is indicated to reverse overdose, yet it is not clear in what quantity fentanyl might have been involved and whether all cases evidenced opioid-induced respiratory depression that would indicate a need for naloxone. Although not directly examined in this study, differences in naloxone administration rates based on suspected xylazine co-involvement may plausibly reflect uncertainty regarding how to respond, considering distinctions between xylazine-associated sedation versus opioid-induced respiratory depression. At the same time, media headlines sometimes describe xylazine as “resistant” to naloxone (e.g., Talbert, 2024), and researchers have posited that statements such as “‘naloxone is ineffective in xylazine intoxication’… propagate misinformation that naloxone should not be used in patients with xylazine-fentanyl overdose” (D’Orazio et al., 2023, p. 1372), potentially impacting law enforcement officers’ beliefs about overdose response.

Differences in naloxone administration rates based on suspected xylazine co-involvement may also reflect the elevated stigma surrounding xylazine use (Bowles et al., 2024) or varying levels of access to naloxone or overdose education in different jurisdictions where xylazine may be more or less prevalent. Moreover, in some portion of the ODIN cases examined in the present study, naloxone might not have been administered because the person was already deceased by the time first responders arrived; 55% of the ODIN-recorded suspected fentanyl-xylazine overdoses were fatal, compared to 27% of the suspected fentanyl-no-xylazine overdoses, but it was unclear what proportion of these deaths occurred before, versus after, a first responder arrived on scene.

### Naloxone dose count differences based on suspected xylazine co-involvement

Results from this study also provided preliminary evidence that when naloxone was administered at the scene of an ODIN-recorded fentanyl overdose, higher numbers of naloxone doses were administered when xylazine was suspected to be co-involved. For instance, 21% of the suspected fentanyl-xylazine overdoses in which naloxone was administered had three or more doses recorded, compared to 10% of the suspected fentanyl-no-xylazine overdoses. This is consistent with findings from qualitative research with medical first responders, harm reduction outreach workers, and persons who use drugs describing a tendency for bystanders and first responders to administer multiple doses of naloxone in cases of xylazine-involved overdose, sometimes without waiting the recommended time between doses (Reed et al., 2025; Quijano et al., 2023). As described in a case series of xylazine-involved overdose events, “patients who would have previously quickly regained consciousness after… naloxone… exhibit 10–30-minute periods of unconsciousness,” (Bufanda et al., 2025, p. 4) potentially leading responders to administer more naloxone even though the person’s breathing may have already been restored by the first dose(s) of naloxone (Reed et al., 2025; Quijano et al., 2023). Naloxone dosage is a topic of ongoing deliberation (Carpenter et al., 2022; Moe et al., 2020) due to the high potency of the synthetic opioids (such as fentanyl) that have inundated US drug supplies, but one of the primary concerns with administering more doses of naloxone than needed is the risk of precipitated withdrawal in persons with physiological dependence on opioids (Hill et al., 2022). In cases of xylazine-fentanyl-involved overdose, therefore, some harm reduction outreach workers and first responders have reported recommending and/or implementing protocol changes such as focusing on the person’s breathing and oxygen levels, rather than their level of consciousness, when determining whether to administer more naloxone, and emphasizing rescue breathing, pulse oximeters, and supplemental oxygen as key to overdose response in addition to naloxone (Bufanda et al., 2025; Friedman et al., 2022; Quijano et al., 2023).

### Other differences based on suspected xylazine co-involvement

Study results highlighted several additional ways in which suspected fentanyl-xylazine ODIN-recorded overdose events differed from fentanyl-no-xylazine overdose events. First, compared to fentanyl-involved overdoses without xylazine suspected, significantly higher proportions of overdoses with xylazine suspected also reportedly involved fentanyl analogs/other synthetic opioids, methamphetamine, and benzodiazepines. This result might reflect additional testing/investigation completed in some overdose events than others, based on factors such as the resources available to different law enforcement agencies and other first responders. At the same time, this finding is consistent with results from analyses of fatal overdoses documenting notable stimulant and benzodiazepine co-involvement in xylazine-fentanyl deaths as well as higher numbers of co-involved drugs in deaths involving xylazine than those not involving xylazine (Bradford et al., 2024; Kariisa et al., 2022; Sibbesen et al., 2023).

Second, among ODIN-recorded fentanyl overdose deaths, a higher proportion of the overdoses with suspected xylazine co-involvement were fatal (55% vs. 27% of the fentanyl-no-xylazine overdoses). The role of xylazine in overdose outcomes is currently unknown (D’Orazio et al., 2023; Hoffman, 2023; Love et al., 2023), and it is unclear to what extent the higher proportion of fatalities in the suspected xylazine-fentanyl ODIN overdoses may reflect fatality risk versus differences in rates of naloxone administration and/or other drug co-involvement. It is also plausible that ODIN-recorded overdose events which were fatal, versus nonfatal, may have received more extensive investigation of drugs involved and/or greater suspicion that additional drugs (such as xylazine) were involved.

Finally, study results indicated that in the subset of ODIN-recorded overdoses in which naloxone was administered and the patient survived, post-naloxone first responder/patient actions differed between suspected fentanyl-xylazine and fentanyl-no-xylazine overdoses. These analyses were limited by the relatively small subset of suspected xylazine-involved cases available yet provided preliminary evidence of several differences. First, patients refused transport in only 5% of the suspected fentanyl-xylazine nonfatal overdoses, compared to 23% of the suspected fentanyl-no-xylazine nonfatal overdoses. Second, patients were transported to the hospital *unconscious* in a higher proportion (14%) of the suspected fentanyl-xylazine nonfatal overdoses than the fentanyl-no-xylazine nonfatal overdoses (6%). These results are consistent with xylazine’s sedative properties and the “long period of post-naloxone sedation in xylazine overdoses [as] a unique clinical feature” (Bufanda et al., 2025, p. 1).

### Implications

Study findings underscore the need for targeted training programs for first responders to address the complexities of xylazine-involved overdoses. Naloxone administration remains critical in suspected xylazine overdoses due to frequent fentanyl co-involvement.

However, education/training should also discuss the risks of over-administering naloxone or administering inappropriately for patients who are heavily sedated rather than experiencing respiratory depression. Evidence-based protocols should guide naloxone use alongside adjunctive measures such as oxygenation and rescue breathing. Practical, scenario-based training—such as simulations of xylazine-fentanyl overdose scenarios—may improve confidence in assessing and managing these cases effectively.

Addressing structural barriers is equally critical, as disparities and deep-seated mistrust between people who use drugs and law enforcement may hinder effective interventions. In Philadelphia, focus groups (Reed et al., 2025) with people who use drugs and harm reduction workers highlighted concerns regarding law enforcement-driven interventions. These focus groups also reflected mixed experiences, some negative, with emergency medical services’ response to xylazine-involved overdoses (Reed et al., 2025). To address these barriers, training programs may integrate trauma-informed care, equipping responders with the skills to build trust and reduce stigma. Collaboration among harm reduction experts, community leaders, first responders, and law enforcement agencies is essential for co-developing protocols that ensure interventions are trauma-informed, culturally sensitive, and aligned with communities’ needs.

### Limitations

ODIN data are not inclusive of, nor representative of, all law-enforcement-attended overdoses in Pennsylvania. Moreover, Philadelphia County (the largest county, one with a particularly diverse population, and often considered the epicenter of xylazine; Friedman et al., 2022) was not included in the study’s analysis, as the Philadelphia Police Department was not listed as an ODIN reporting agency at the time of the study. All ODIN measures, including drug involvement, are based on first-responder assessment/report. ODIN reporters use a variety of methods to identify suspected drugs involved (including drug field testing, interviews with those at the scene, and investigation of drugs and paraphernalia present; Barboza & Angulski, 2020), and it is plausible that responders may be more likely to suspect and/or report more drugs in fatal than non-fatal events and that investigation practices and resources may differ between agencies.

ODIN data may over-report historically common drugs (e.g., heroin) and under-report emerging drugs such as xylazine. It is unclear to what extent ODIN xylazine underreporting may compare to the level of xylazine underreporting in mortality data and nonfatal overdose surveillance data (attributed to limited testing; Spencer et al., 2023; Korona-Bailey et al., 2023) or the level of discrepancy between residue xylazine test strip results and laboratory-based testing (Thompson et al., 2024). At the same time, first responders’ assessment of drug involvement (based on the information and investigation resources available at the scene and their prior experiences/training/perceptions) is what shapes their decisions regarding how to respond, since full toxicological testing results would not be available at the time of overdose response to inform in-the-moment decisions.

The study’s analyses were limited by the modest sample sizes of suspected xylazine-fentanyl overdose events recorded in the data. Analyses of naloxone dose counts did not include information on method of administration (not available in the data) or exact dose size (due to the level of missing data on the dose milligram measures in the dataset). Finally, other potentially relevant measures were not available in the data source, including individuals’ underlying health status and opioid tolerance levels, symptomatology, life circumstances, quantity of drugs consumed, other emergency response interventions received, and whether death (in the subset of fatal overdoses) occurred before or after a first responder arrived. We were also unable to assess first responders’ specific job position, training history, the resources and procedures of each local area’s emergency system, and the exact information (e.g., interviews, evidence on scene, drug field testing) available to determine which drugs were suspected in any individual overdose.

## Conclusions

Study results provided evidence of relatively lower rates of naloxone administration in suspected fentanyl-xylazine-involved overdoses (versus fentanyl-no-xylazine overdoses) recorded by ODIN-participating law enforcement/first responders in Pennsylvania outside of Philadelphia. While naloxone does not directly reverse the effects of xylazine, it remains effective in reversing the effects of the fentanyl with which xylazine is often mixed (Gupta et al., 2023). As such, results of the present study underscore the relevance of providing both naloxone and specific training regarding xylazine-fentanyl overdose to first responders, incorporating best practices identified by medical and harm reduction outreach workers with experience responding to xylazine-involved overdoses (e.g., Bufanda et al., 2025; Quijano et al., 2023) and the perspectives and concerns of persons with lived experience (e.g., Reed et al., 2025). Public health and harm reduction programs play a vital role in disseminating evidence-based information on the local drug supply, naloxone, and rescue breathing and other lifesaving care for individuals with suspected overdoses involving fentanyl and xylazine. Study findings underscore the importance of making low-threshold harm reduction and drug checking services widely available to people who use drugs while also equipping first responders and likely bystanders with the tools and training to recognize and manage the distinct challenges associated with xylazine-polysubstance overdoses.

## Compliance and Ethical Standards

The study was classified as “exempt” by the Arizona State University Institutional Review Board on July 18, 2024, based on Federal Regulation 45CFR46 (4). The study consisted of analyses of publicly-available de-identified data only.

## Data Availability

The data used in this study were obtained from a publicly-available dataset at: https://data.pa.gov/Opioid-Related/Overdose-Information-Network-Data-CY-January-2018-/hbkk-dwy3/about_data

---

**Supplemental Table S1:** Steps taken to identify unique records in the Overdose Information Network (ODIN) dataset and merge data on multiple drugs suspected to be involved in each overdose.

| Step | Rationale/Details |
| --- | --- |
| Use ODIN “victim ID” number to identify unique “victims” within any given overdose incident reported | Some overdose events with the same ODIN “incident ID” include different persons (with different “victim ID” numbers) who suffered an overdose together. |
| Examine records with the same “victim ID” number to determine whether they all correspond to the same “incident ID” and date/time | Records with the same “victim ID” but different “incident ID” and date/time would indicate the same person experiencing different overdose incidents at different times recorded in the ODIN.<br>No records with the same “victim ID” but different “incident IDs” were located, suggesting that any repeat overdoses in the same victim were not identifiable, due to assignment of a new victim ID each incident. |
| Examine and compare the study measures (suspected drug, sex, age, race, ethnicity, county of occurrence, year, suspected drug, naloxone administration, naloxone dose, survival outcome, and post-naloxone action [“revive action description”]) between records with the same victim ID/incident ID combination | The ODIN includes multiple records with the same victim ID/incident ID combination.<br>Examination of such multiple records revealed that the sex, age, race, ethnicity, county of occurrence, date/time, naloxone administration, survival outcome, and post-naloxone action were consistent/the same between all records with the same victim ID/incident ID combination.<br>In contrast, the “suspected drug” field differed between multiple records.<br>The naloxone dose quantity field also differed in a very low proportion of records. |
| Merge records by victim ID | Each row in the original ODIN Excel file (as downloaded from the Open PA website) can accommodate only one drug type response, due to only one “suspected drug” field available in each row. Thus, multiple rows for the same victim and incident are used to reflect multiple drug types suspected to be involved in any given person’s overdose incident.<br>Merging based on victim ID (since there were no instances of the same victim ID with different incident IDs) allows us to maintain only one unique record for each victim while maintaining multiple fields for the “suspected drugs.” All other measures examined in the study (except naloxone dose quantity, mentioned below) were consistent within each victim ID.<br>In analyses of naloxone dose quantity, we retained only the records with consistent data on naloxone dose quantity (resulting in the loss of less than 2% of records in these analyses). |

**Supplemental Table S2.**
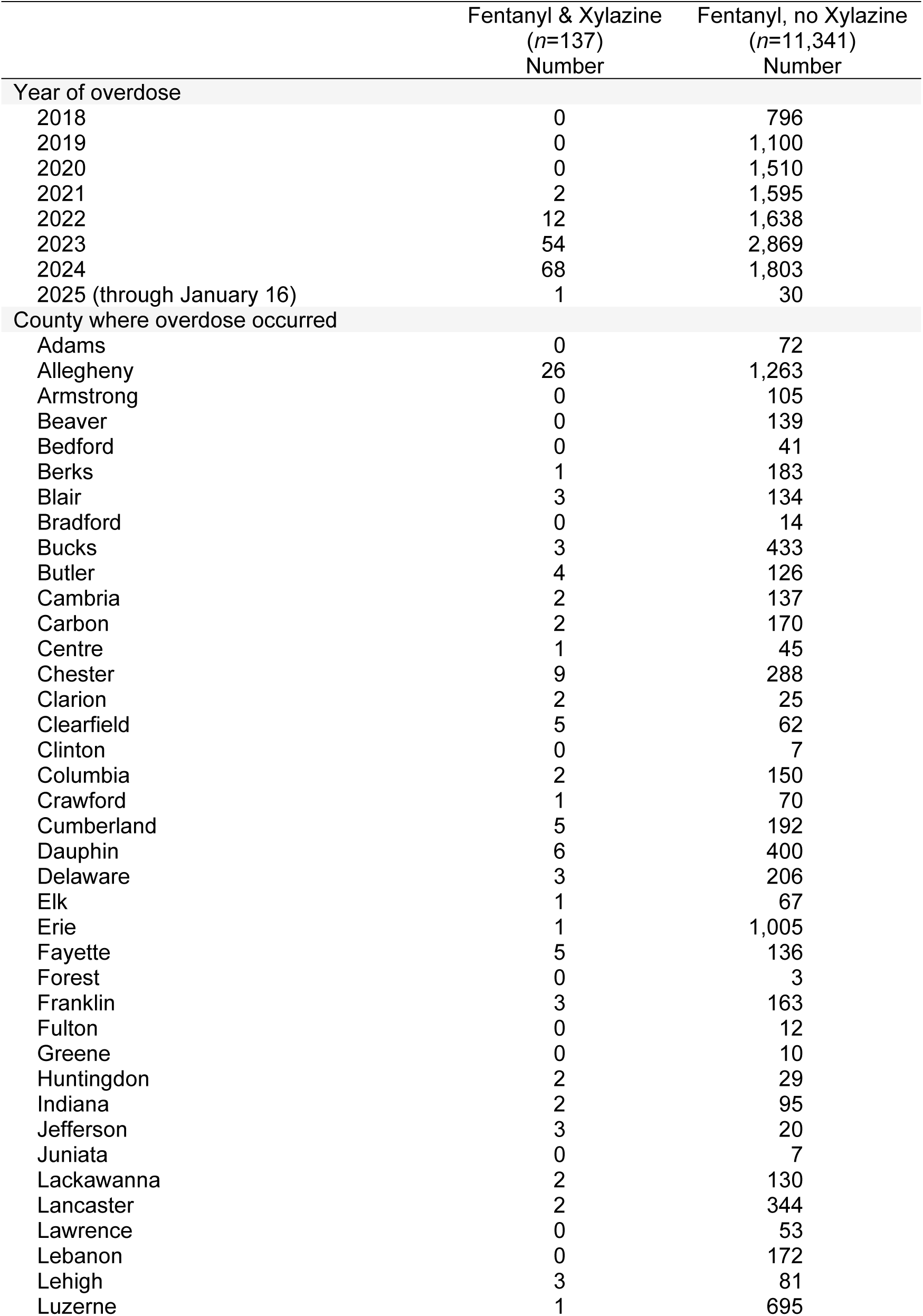
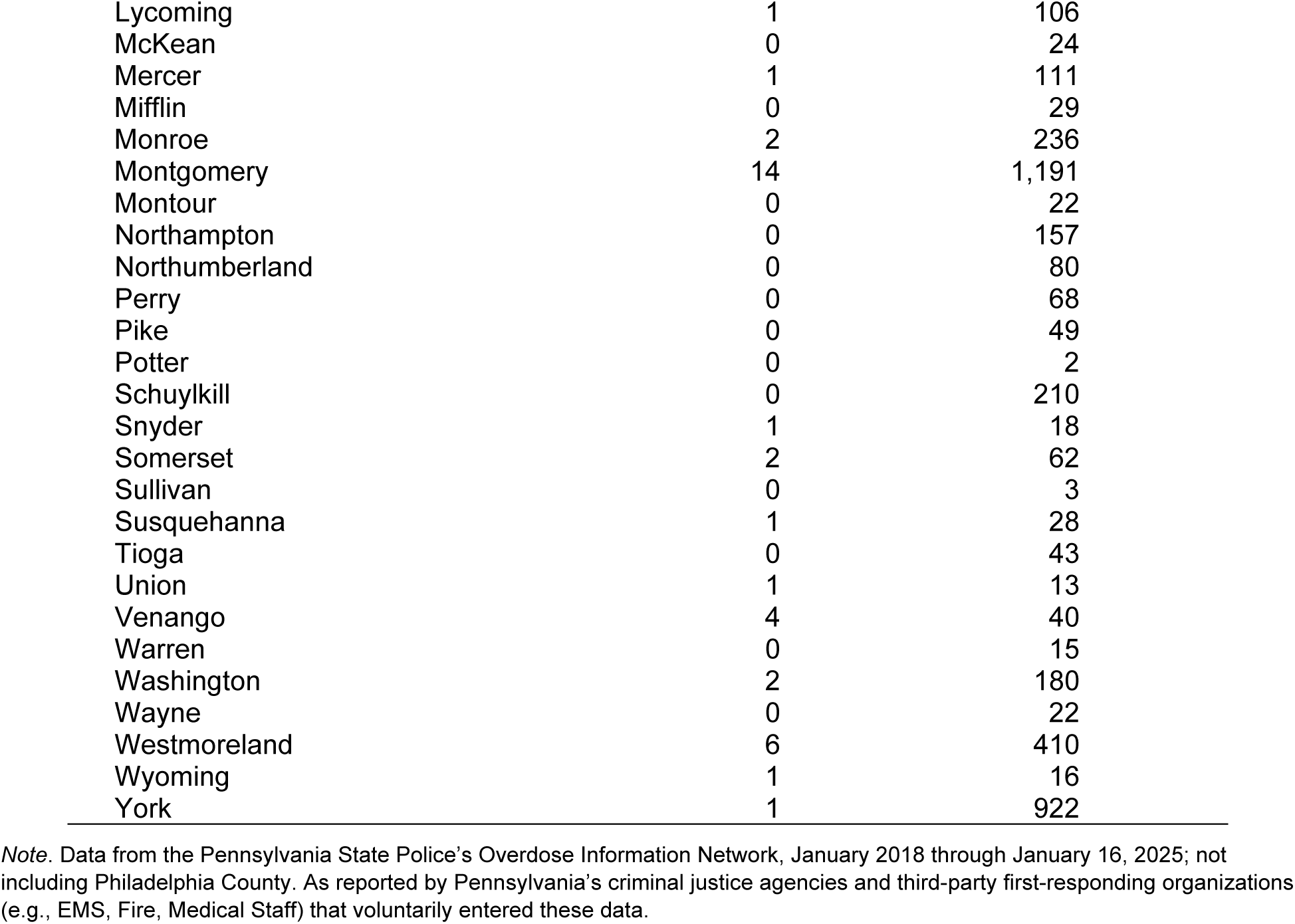
Number of suspected overdose events reportedly involving fentanyl and xylazine versus fentanyl and no xylazine, by county and year, as recorded in the Pennsylvania Overdose Information Network, January 2018-January 16, 2025.

**Supplemental Table S3.** Characteristics of suspected overdose events reportedly involving fentanyl and xylazine versus fentanyl and no xylazine, as recorded in the Pennsylvania Overdose Information Network, January 2018-January 16, 2025.

|  | Fentanyl & Xylazine<br>n=137<br>% | Fentanyl, no Xylazine<br>n=11,341<br>% | P value<sup>§</sup> |
| --- | --- | --- | --- |
| Age, years |  |  | 0.080 |
| 0-9 | 0.7 | 0.3 |  |
| 10-19 | 0.0 | 1.4 |  |
| 20-29 | 18.3 | 23.3 |  |
| 30-39 | 35.8 | 38.5 |  |
| 40-49 | 31.4 | 21.0 |  |
| 50-59 | 9.5 | 10.7 |  |
| 60-69 | 3.7 | 4.4 |  |
| 70+ | 0.7 | 0.5 |  |
| Sex |  |  | 0.344 |
| Male | 65.7 | 71.1 |  |
| Female | 34.3 | 28.7 |  |
| Unknown | 0.0 | 0.2 |  |
| Race/ethnicity |  |  | 0.308 |
| NH White | 83.2 | 80.5 |  |
| NH Black | 7.3 | 9.0 |  |
| Hispanic | 5.1 | 6.0 |  |
| NH AI/AN | 0.7 | 0.1 |  |
| NH Asian | 0.0 | 0.2 |  |
| Unknown | 3.7 | 4.2 |  |
| Suspected drug involved in the overdose<sup>1</sup>: |  |  |  |
| Heroin | 46.0 | 66.3 | <0.001 |
| Carfentanil | 10.2 | 2.2 | <0.001 |
| Other synthetic opioid/fentanyl analog | 20.4 | 7.6 | <0.001 |
| Pharmaceutical opioid | 1.5 | 3.0 | 0.444 |
| Methamphetamine | 20.4 | 7.0 | <0.001 |
| Cocaine/crack | 10.2 | 9.1 | 0.652 |
| Benzodiazepines | 5.1 | 2.4 | 0.046 |
| Alcohol | 4.4 | 2.5 | 0.156 |
| Naloxone administered? |  |  | <0.001 |
| Yes | 46.0 | 67.3 |  |
| No | 54.0 | 32.7 |  |
| Survived? |  |  | <0.001 |
| Yes | 41.6 | 68.9 |  |
| No | 54.7 | 27.0 |  |
| Unknown | 3.7 | 4.2 |  |

Note. Data from the Pennsylvania State Police's Overdose Information Network, January 2018 through January 16, 2025, not including Philadelphia County. <sup>1</sup>As reported by Pennsylvania's criminal justice agencies and third-party first-responding organizations (e.g., EMS, Fire, Medical Staff) that voluntarily entered these data. <sup>§</sup>P values calculated based on Fisher's Exact Tests. Abbreviations: NH, Non-Hispanic; AI/AN, American Indian/Alaska Native.

**Supplemental Table S4.** Sensitivity analysis results from binomial logistic regression analysis predicting naloxone administration, based on xylazine involvement and selected covariates, among suspected overdoses involving any opioid or fentanyl-related overdoses (2022-2025) reported in the Pennsylvania Overdose Information Network.

|  | Overdoses Involving Any Opioid,<br>2018-2025<sup>a</sup> | Fentanyl Overdoses,<br>2022-2025<sup>a</sup> |
| --- | --- | --- |
|  | Outcome:<br>Naloxone<br>Administration<br>*n*=25,799 | Outcome:<br>Naloxone<br>Administration<br>*n*=6,188 |
|  | AOR [95% CI] | AOR [95% CI] |
| Xylazine co-involved | **0.45\*\*\*** [0.32, 0.63] | **0.40\*\*\*** [0.27, 0.57] |
|  | AME [95% CI] | AME [95% CI] |
| Average Marginal Effect<br>(AME), xylazine [95% CI] | **-0.16\*\*\*** [-0.23, -0.09] | **-0.19\*\*\*** [-0.27, -0.12] |

*Note.* Naloxone administration denotes if naloxone was administered to the individual who overdosed. Data are from January 2018 through January 16, 2025; data do not include Philadelphia County. Each regression model includes the following covariates: age, sex, race/ethnicity, drugs co-involved in the overdose (heroin, carfentanil, other synthetic opioid/fentanyl analog, pharmaceutical opioids, methamphetamine, cocaine/crack, benzodiazepines, and alcohol), county rurality, and year-fixed effects. <sup>a</sup>Through January 16, 2025. Average Marginal Effects (AME) denote the proportional change in the outcome when xylazine changes from “not suspected to be involved” to “suspected to be involved in the overdose.” Estimates in bold highlight statistical significance (two-tailed): \**p* < .05, \*\**p* < .01, \*\*\**p* < .001. *Abbreviations:* AOR, adjusted odds ratio; CI, confidence interval.

**Supplemental Table S5.** Survival outcomes based on the number of naloxone doses administered, for 7,550 ODIN-recorded suspected fentanyl-related overdoses in which naloxone was administered, January 2018-January 16, 2025.

| Number of Naloxone Doses Administered | <u>Did the person survive the overdose event?</u> |  |  |
| --- | --- | --- | --- |
|  | No | Yes | Unknown/Missing |
| 1 dose | 9.6 | 85.7 | 4.7 |
| 2 doses | 8.9 | 85.7 | 5.3 |
| 3+ doses | 11.2 | 84.8 | 4.0 |

Notes. Data are from the Pennsylvania Overdose Information Network (ODIN), January 2018 through January 16, 2025; data do not include Philadelphia County. Pearson $\chi^2(4) = 5.63$ , $p=0.228$ ; Fisher's exact, $p=0.235$ .

## References

Barboza G. E., Angulski, K. 2020. A descriptive study of racial and ethnic differences of drug overdoses and naloxone administration in Pennsylvania. International Journal of Drug Policy 78:102718. 10.1016/j.drugpo.2020.102718

Bowles J. M., Copulsky, E. C., Reed, M. K. 2024. Media framing xylazine as a “zombie drug” is amplifying stigma onto people who use drugs. International Journal of Drug Policy 125:104338. 10.1016/j.drugpo.2024.104338

Bradford W., Figgatt, M., Scott, K. S., Marshall, S., Eaton, E. F., Dye, D. W. 2024. Xylazine co-occurrence with illicit fentanyl is a growing threat in the Deep South: A retrospective study of decedent data. Harm Reduction Journal 21(1):46. doi:10.1186/s12954-024-00959-2

Bufanda, L. P., Montoya, A. G., Carrillo, B. T., Tejeda, M. A. G., Segovia, L. A., Calderón-Villarreal, A., & Friedman, J. R. 2025. Managing xylazine-involved overdoses in a community harm reduction setting: lessons from Tijuana, Mexico. Harm Reduction Journal 22:2. 10.1186/s12954-024-01143-2

Cano M., Daniulaityte R, Marsiglia F. Xylazine in overdose deaths and forensic drug reports in US States, 2019-2022. 2024. JAMA Network Open 7(1):e2350630. doi:10.1001/jamanetworkopen.2023.50630

Cano, M., Jones, A., Silverstein, S. M., Daniulaityte, R., & LoVecchio, F. 2025. Naloxone administration and survival in overdoses involving opioids and stimulants: An analysis of law enforcement data from 63 Pennsylvania counties. International Journal of Drug Policy, 135: 104678. 10.1016/j.drugpo.2024.104678

Carpenter, J., Murray, B. P., Atti, S., Moran, T. P., Yancey, A., & Morgan, B. 2020. Naloxone dosing after opioid overdose in the era of illicitly manufactured fentanyl. Journal of Medical Toxicology 16:41–48. 10.1007/s13181-019-00735-w

Choi, S., Irwin, M. R., Kiyatkin, E. A. 2023. Xylazine effects on opioid-induced brain hypoxia. Psychopharmacology (Berl) 240(7):1561–1571. 10.1007/s00213-023-06390-y

Copeland, C. S., Rice, K., Rock, K. L., et al. 2024. Broad evidence of xylazine in the UK illicit drug market beyond heroin supplies: Triangulating from toxicology, drug-testing and law enforcement. Addiction 119(7):1301–1309. doi:10.1111/add.16466

D’Orazio J, Nelson L, Perrone J, Wightman R, Haroz R. 2023. Xylazine adulteration of the heroin-fentanyl drug supply: A narrative review. Annals of Internal Medicine 176(10):1370–1376. doi:10.7326/M23-2001

Delcher, C., Quesinberry, D., Torabi, S., et al. 2024. Wastewater surveillance for xylazine in Kentucky. AJPM Focus 3(3):100203. doi:10.1016/j.focus.2024.100203

Del Pozo, B. 2022. Reducing the iatrogenesis of police overdose response: Time is of the essence. American Journal of Public Health 112(9): 1236–1238. 10.2105/AJPH.2022.306987

Doe-Simkins, M., El-Sabawi, T., & Carroll, J. J. 2022. Whose concerns? It’s time to adjust the lens of research on police-involved overdose response. American Journal of Public Health, 112(9):1239–1241. 10.2105/AJPH.2022.306988

Fairbairn, N., Coffin, P. O., & Walley, A. Y. 2017. Naloxone for heroin, prescription opioid, and illicitly made fentanyl overdoses: Challenges and innovations responding to a dynamic epidemic. International Journal of Drug Policy 46:172–179. 10.1016/j.drugpo.2017.06.005

Friedman, J. R., Montero, F., Bourgois, P., et al. 2022. Xylazine spreads across the US: A growing component of the increasingly synthetic and polysubstance overdose crisis. Drug and Alcohol Dependence 233:109380. doi: 10.1016/j.drugalcdep.2022.109380

Friedman, J. R., Montoya, A. G., Ruiz, C., et al. 2024. The detection of xylazine in Tijuana, Mexico: Triangulating drug checking and clinical urine testing data. Preprint. medRxiv. doi:10.1101/2024.08.19.24312273

Gupta R., Holtgrave, D. R., Ashburn, M. A., 2023. Xylazine - Medical and Public Health Imperatives. The New England Journal of Medicine 388(24):2209–2212. doi: 10.1056/NEJMp2303120

Hill, L. G., Zagorski, C. M., & Loera, L. J. 2021. Increasingly powerful opioid antagonists are not necessary. International Journal on Drug Policy 99:103457. 10.1016/j.drugpo.2021.103457

Hoffman, R. S. 2023. Closing the xylazine knowledge gap. Clinical Toxicology 61(12):1013–1016. 10.1080/15563650.2023.2294619

Holmes, L. M., & King, B. H. 2023. County-level variation in synthetic opioid and heroin overdose incidents in Pennsylvania during the COVID-19 pandemic. Applied Geography 155:102977. 10.1016/j.apgeog.2023.102977

Holmes, L. M., Rishworth, A., King, B. H. 2022. Disparities in opioid overdose survival and naloxone administration in Pennsylvania. Drug and Alcohol Dependence 238:109555. doi:10.1016/j.drugalcdep.2022.109555

Jacoby, J. L., Crowley, L. M., Cannon, R. D., Weaver, K. D., Henry-Morrow, T. K., Henry, K. A., Kayne, A. N., Urban, C. E., Gyory, R. A., & McCarthy, J. F. 2020. Pennsylvania law enforcement use of Narcan. The American journal of Emergency Medicine 38(9):1944–1946. 10.1016/j.ajem.2020.01.051

Kariisa, M., O’Donnell, J., Kumar, S., Mattson, C. L., Goldberger, B. A. 2022. Illicitly manufactured fentanyl-involved overdose deaths with detected xylazine - United States, January 2019-June 2022. Morbidity and Mortality Weekly Report 72(26):721–727. 10.15585/mmwr.mm7226a4

King, B., Patel, R., & Rishworth, A. 2021. Assessing the relationships between COVID-19 stay-at-home orders and opioid overdoses in the State of Pennsylvania. Journal of Drug Issues 51(4):648–660. 10.1177/00220426211006362

Koester, S., Mueller, S. R., Raville, L., Langegger, S., & Binswanger, I. A. 2017. Why are some people who have received overdose education and naloxone reticent to call Emergency Medical Services in the event of overdose? International Journal of Drug Policy 48:115–124. 10.1016/j.drugpo.2017.06.008

Korona-Bailey, J., Onyango, E., Hall, K. F., Jayasundara, J., Mukhopadhyay, S. 2023. Xylazine-involved fatal and nonfatal drug overdoses in Tennessee from 2019 to 2022. JAMA Network Open 6(7):e2324001. doi:10.1001/jamanetworkopen.2023.24001

Love, J. S., Levine, M., Aldy, K., et al. 2023. Opioid overdoses involving xylazine in emergency department patients: A multicenter study. Clinical Toxicology 61(3):173–180. 10.1080/15563650.2022.2159427

Moe, J., Godwin, J., Purssell, R., O’Sullivan, F., Hau, J. P., Purssell, E. et al. 2020. Naloxone dosing in the era of ultra-potent opioid overdoses: A systematic review. Canadian Journal of Emergency Medicine 22(2):178–186. doi:10.1017/cem.2019.471

Pennsylvania State Police. 2024. Overdose Information Network Data CY January 2018-Current Monthly County State Police. https://data.pa.gov/Opioid-Related/Overdose-Information-Network-Data-CY-January-2018-/hbkk-dwy3/about_data

Quijano, T., Crowell, J., Eggert, K., Clark, K., Alexander, M., Grau, L., & Heimer, R. 2023. Xylazine in the drug supply: Emerging threats and lessons learned in areas with high levels of adulteration. International Journal of Drug Policy 120:104154. 10.1016/j.drugpo.2023.104154

Reed, M. K., Esteves Camacho, T., Olson, R., Grover, Z., Rapoza, T., & Larson, M. J. 2025. Xylazine’s impacts on the community in Philadelphia: Perspectives of people who use opioids and harm reduction workers. Substance Use & Misuse, 60: 100–107.

Rhoads, D. 2019. Pennsylvania Overdose Information Network (ODIN): Executive summary. https://www.nascio.org/wp-content/uploads/2020/09/PA-Data-Mgmt-OverdoseInformation-Network-NASCIO-2019-FINAL.pdf

S.B. 1152, P.L. 2158. Act No. 158 of 2022. Overdose mapping act-enactment. Pennsylvania General Assembly. https://www.legis.state.pa.us/cfdocs/legis/li/uconsCheck.cfm?yr=2022&sessInd=0&act=158#:~:text=OVERDOSE%20MAPPING%20ACT%20%2D%20ENACTMENT&text=Establishing%20the%20Overdose%20Information%20Network,on%20the%20Pennsylvania%20State%20Police.

Sibbesen, J., Abate, M. A., Dai, Z., et al. 2023. Characteristics of xylazine-related deaths in West Virginia-Xylazine-related deaths. The American Journal on Addictions 32(3):309–313. doi:10.1111/ajad.13365

Spencer, M. R., Cisewski, J. A., Warner, M., Garnett, M. F. 2023. Drug overdose deaths involving xylazine: United States, 2018–2021. Vital Statistics Rapid Release; no 30. 10.15620/cdc:129519

Talbert, B. 2024. Jefferson Co. Coroner Warns of Narcan Resistant Drug Killing People. WBRC News, September 20. Accessed January 26, 2025. https://www.wbrc.com/2024/09/20/jefferson-co-coroner-warns-narcan-resistant-drug-killing-people/

Thomas, C., Mondal, A., Levitas, M., Widman, C., Lemberg, B. 2024. Trends in tranq: Prevalence of xylazine in oral fluid toxicology in Michigan, Ohio, and Indiana. Preprint. Research Square. 10.21203/rs.3.rs-4669492/v1

Thompson, E., Tardif, J., Ujeneza, M., et al. 2024. Pilot findings on the real-world performance of xylazine test strips for drug residue testing and the importance of secondary testing methods. Drug and Alcohol Dependence Reports 11:100241. 10.1016/j.dadr.2024.100241

Wagner, K. D., Harding, R. W., Kelley, R., Labus, B., Verdugo, S. R., Copulsky, E. et al. 2019. Post-overdose interventions triggered by calling 911: Centering the perspectives of people who use drugs (PWUDs). PLOS One 14(10):e0223823. 10.1371/journal.pone.0223823

Wu, P. E., Austin, E. 2024. Xylazine in the illicit opioid supply. Canadian Medical Association Journal 196(4):E133. doi:10.1503/cmaj.231603

Zagorski, C. M., Hosey, R. A., Moraff, C., et al. 2023. Reducing the harms of xylazine: Clinical approaches, research deficits, and public health context. Harm Reduction Journal 20(1):170. 10.1186/s12954-023-00879-7

Zhu, D. T. 2023. Public health impact and harm reduction implications of xylazine-involved overdoses: A narrative review. Harm Reduction Journal 20(1):131. 10.1186/s12954-023-00867-x

